# Stakeholder Perspectives on the Adaptability of Hospital Drug Formularies to Disease Patterns: A Modified Q-Methodology Study in Vietnam

**DOI:** 10.64898/2025.12.18.25342605

**Authors:** Mai-Hien Chuc, Thi-Thuan Tran, Hai-Anh Ha

## Abstract

**Background:** Hospital drug formularies function within Complex Adaptive Systems (CAS), where the alignment between drug supply and disease patterns is critical for patient care and cost-effectiveness. However, measuring this adaptability is challenging due to conflicting priorities among stakeholders and a lack of standardized assessment tools.

**Objectives:** This study aims to analyze the perspectives of three key stakeholder groups-clinical providers, non-clinical service providers, and policymakers-to develop a consensus-based set of indicators for monitoring the adaptability of drug formularies in Vietnam.

**Methods:** We employed a modified Q-methodology approach involving 69 experts (20 non-clinical, 28 clinical, and 21 policymakers). A Q-set of 92 indicators across 9 criteria was developed. Participants evaluated these indicators using a 5-point Likert scale to determine levels of consensus and discordance, rather than a forced-distribution sort, to assess the absolute importance of adaptability metrics.

**Results:** The analysis revealed high consensus across all groups regarding the importance of “Usage adaptation” and “Storage adaptation”. However, significant discordance was observed in criteria related to “Ensuring drug availability,” “Appropriate procurement” and “Outputs and outcomes”. Notably, non-clinical providers prioritized proactive ordering, whereas clinical experts emphasized output results.

**Conclusion:** The study proposes a management framework that monitors drug list adaptability across four levels: non-adaptive, passive, active, and advanced adaptive.

Successful adaptability requires enhanced collaboration between specialized teams and a shift from efficiency-based to resilience-based performance metrics.

## 1. INTRODUCTION

Hospital drug formularies and supply chain operations are primary agents within the healthcare system-a complex adaptive system (CAS) [1], designed to optimize patient care through the safe, appropriate, and cost-effective use of medicines [2]. As a CAS, the healthcare environment involves multiple subsystems of interconnected actors [3], including physicians, patients, suppliers, and regulators, whose relationships co-evolve and require careful management [4]. Consequently, decision-making in this context is not linear but relies on the storage, retrieval, and processing of knowledge from various stakeholders to support adaptive decisions [5].

The management of these systems faces significant challenges. The pharmaceutical supply chain is heavily influenced not only by demand fluctuations but also by rigid health system regulations and the inherent risks of drug supply operations [6]. In the public sector, the lack of internal cost pressure often leads to inefficiencies, such as slow demand forecasting and poor coordination between supply and demand [1], [7]. Furthermore, conventional performance metrics typically focus on efficiency, overlooking critical factors like sustainability, flexibility, and responsiveness, key components of the emerging Industry 5.0 paradigm [8], [9].

While recent literature has emphasized supply chain resilience (the ability to recover from disruptions) [10], [11], there is a distinction between resilience and adaptability [12]. Adaptability refers to the capacity to evolve continuously without disruption, ensuring the availability of medicines through proactive adjustments [13]. Currently, there is a lack of comprehensive tools to monitor and evaluate the specific adaptability of drug formularies to changing disease patterns. Existing frameworks often suffer from limited stakeholder engagement, which undermines the adaptive capacity of health systems [4], [14], [15].

Since stakeholders hold varying and often conflicting perspectives on what constitutes “value” or “efficiency” in drug supply, it is essential to capture these diverse viewpoints [14]. A robust evaluation framework must be flexible enough to incorporate these conflicts and guide decision-making toward a balance between maximizing benefits and minimizing adverse impacts [16].

Therefore, this study aims to: (1) analyze stakeholder perspectives on the adaptability of drug formularies to disease patterns using a structured survey method based on Q-methodology; and (2) provide management implications for improving supply chain adaptability. By identifying areas of consensus and discordance among clinical, non-clinical, and policy experts, this research contributes to the development of a unified set of indicators for monitoring hospital drug formulary performance in Vietnam.

## 2. MATERIALS AND METHODS

### 2.1. Study design

This study employed a modified Q-methodology approach, utilizing a cross-sectional survey design. While traditional Q-methodology relies on a forced-distribution sorting technique to identify distinct viewpoints (factors), this study adapted the method by applying a Likert-scale evaluation to the Q-set items. This hybrid design allowed for the rigorous development of adaptability indicators (characteristic of Q-methodology) while enabling the assessment of absolute importance and consensus levels across stakeholder groups through quantitative statistical analysis.

### 2.2. Development of the Q-set (research instrument)

The “Q-set” (the collection of statements) was constructed based on a comprehensive literature review and expert consultations regarding hospital drug formularies and supply chain adaptability. The development process involved two phases of expert consultation to ensure content validity. Initially, a preliminary set of indicators was distributed to experts for evaluation and open-ended feedback (**S1 Appendix**). Based on the feedback and quantitative scoring from this first round, indicators with low agreement (mean score < 3.5) were removed, and new indicators proposed by experts were added. The refined instrument was then finalized for the official consensus rating (**S2 Appendix**).

The final Q-set comprised 92 indicators categorized into 09 key criteria, covering critical aspects including: (1) Adaptive workforce (staffing); (2) Ensuring drug availability requirements; (3); Methods for drug needs assessment; (4) Ensuring legal compliance, (5) Appropriate procurement (ordering); (6) Usage adaptation; (7) Storage adaptation; (8) Risk management; (9) Appropriate outputs and outcomes.

### 2.3. Participant selection (P-set)

A purposive sampling strategy was used to select the “P-set” (participants) to ensure diverse representation of the healthcare supply chain ecosystem. The study involved 69 experts (N=69) divided into three distinct stakeholder groups:

- Non-clinical service providers (NC): N = 20 (pharmaceutical companies, distributors).
- Clinical service providers (CP): N = 28 (hospital pharmacists, doctors, clinical managers).
- Policymakers (PM): N = 21 (health authorities, regulatory bodies).

### 2.4. Data collection procedure

This study engaged experts solely in their professional capacity to provide opinions on management indicators, involving no clinical intervention, patient data, or biological samples. Therefore, formal ethical approval was waived in accordance with local regulations for non-biomedical administrative research. However, the study adhered strictly to the ethical principles of the Declaration of Helsinki.

The data collection was conducted from September 1^st^ 2024 to October 1^st^ 2024 (round 1) and from November 15^th^ 2024 to December 31^st^ 2024 (round 2). Informed consent was obtained from all participants. The survey cover letter clearly explained the study’s purpose, anonymity, and voluntary nature. Returning the completed questionnaire was considered as implied consent to participate.

The data collection followed a rigorous three-step process (**Figure 1**):

- Q-set construction: Establishing the 92 statements (as described above).
- P-set recruitment: Selecting the 69 experts.
- Modified Q-sort (rating): Instead of sorting cards into a forced distribution curve, participants were asked to rate their agreement with each of the 92 statements on a 5-point Likert scale (1 = Strongly disagree to 5 = Strongly agree). This modification allowed experts to express the absolute importance of multiple high-priority indicators simultaneously, which is often restricted in traditional Q-sorting. Data were entered and independently cross-checked three times to ensure accuracy before statistical processing.

**Figure 1.**
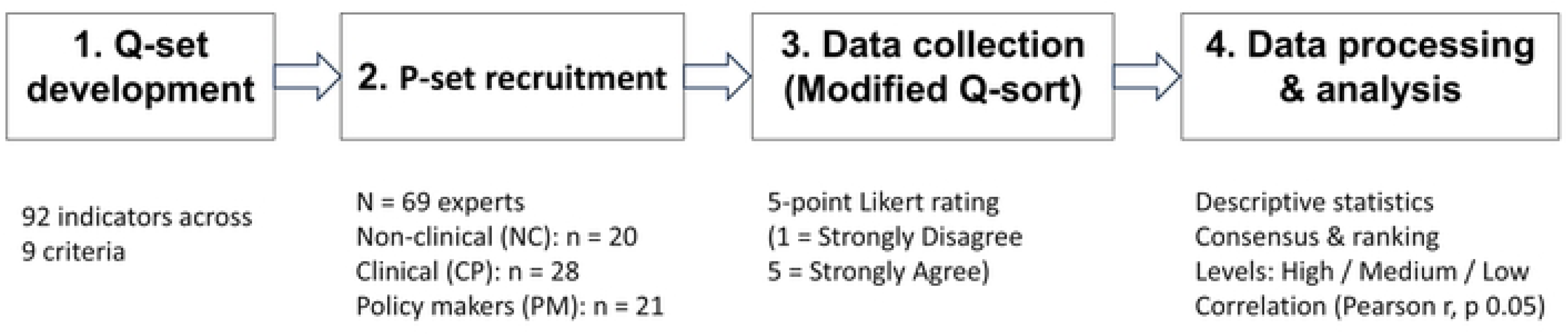
Workflow for developing and evaluating adaptability indicators using a modified Q-methodology approach.

### 2.5. Statistical analysis

Data were processed using Microsoft Excel. The analysis focused on identifying consensus and discordance among the three stakeholder groups.

- Descriptive statistics: Frequencies and mean scores were calculated for each indicator. Normality of the data distribution was assessed using Z-scores for skewness and kurtosis. The results indicated that the data approximated a normal distribution, with Z-values falling within the acceptable range of ±1.96 (at 5% significance level) or ±2.58 (at 10% significance level).
- Consensus and ranking: Indicators were ranked based on mean scores. Consensus was determined by comparing the rankings across the three groups. Items were classified into three adaptability levels based on their scores: High, Medium, and Low.
- Correlation analysis: Pearson correlation coefficients (r) were calculated to examine the relationship between stakeholder perspectives. The correlation was considered statistically significant if the p-value was ≤ 0.05. A strong positive correlation (r close to 1 with p ≤ 0.05) indicated high agreement between groups, whereas a weak correlation or p > 0.05 indicated divergence in perspectives.

## 3. RESULTS

3.1. Overall adaptability ratings and consensus levels

The assessment of adaptability across the nine criteria revealed generally positive perceptions among stakeholders, with seven out of nine criteria achieving a mean score above the “Agree” threshold (Mean > 4.0). The criteria “Methods for Determining Drug

Needs” (3.96) and “Adaptability Outputs/Results” (3.91) scored slightly lower but remained well above the acceptable threshold of 3.5. **Table 1** presents the mean scores and the classification of consensus versus discordance among the three stakeholder groups.

**Table 1.** Perspectives on the measurability of adaptability Notes: Mean score is reported as mean ± SD on a 5-point Likert scale (1–5). NC = non-clinical service providers; CP = clinical service providers; PM = policy makers. H/M/L = High/Medium/Low.

| Criterion | Mean score | CP | NC | PM | Consensus |
| --- | --- | --- | --- | --- | --- |
| Adaptive workforce (staffing) | 4.17 ± 0.86 | M | M | M | Consensus |
| Ensuring drug availability requirements | 4.07 ± 0.88 | L | M | L | Disagreement |
| Methods for drug needs assessment | 3.96 ± 0.84 | L | L | L | Consensus |
| Ensuring legal compliance | 4.14 ± 0.84 | M | M | M | Consensus |
| Appropriate procurement (ordering) | 4.10 ± 0.87 | M | H | M | Disagreement |
| Usage adaptation | 4.37 ± 0.72 | H | H | H | Consensus |
| Storage adaptation | 4.27 ± 0.80 | H | H | H | Consensus |
| Risk management | 4.00 ± 0.81 | L | L | L | Consensus |
| Appropriate outputs and outcomes | 3.91 ± 0.76 | M | L | L | Disagreement |

The analysis identified distinct levels of consensus. The highest agreement was observed in “Usage Adaptation” and “Storage Adaptation”. This reflects that adaptability is most evident and measurable during the operational stages of drug usage and storage, where reserve calculations and capacity planning are concrete. “Adaptive workforce” and “Ensuring legal compliance” showed medium consensus. Stakeholders recognized these as foundational elements for any adaptive activity. Disagreement was noted in three criteria: “ Ensuring drug availability requirements”, “Appropriate procurement” and “Appropriate outputs and outcomes”. While the divergence was not extreme, it highlights varying strategic priorities between providers and policymakers.

### 3.2. Divergence in stakeholder perspectives

A deeper analysis of the discordant criteria reveals specific structural conflicts in perspectives (**Figure 2**). Regrading ordering adaptation, the non-clinical service providers (NC) rated “Appropriate procurement” significantly higher (rank 1) compared to the clinical (CP) and policy maker (PM) groups, who ranked it at a medium level (rank 5). This discrepancy suggests that suppliers view proactive ordering as a critical active measure for supply security, whereas hospitals and policymakers tend to view successful ordering as an expected standard duty of the supplier rather than an adaptive achievement.

**Figure 2.**
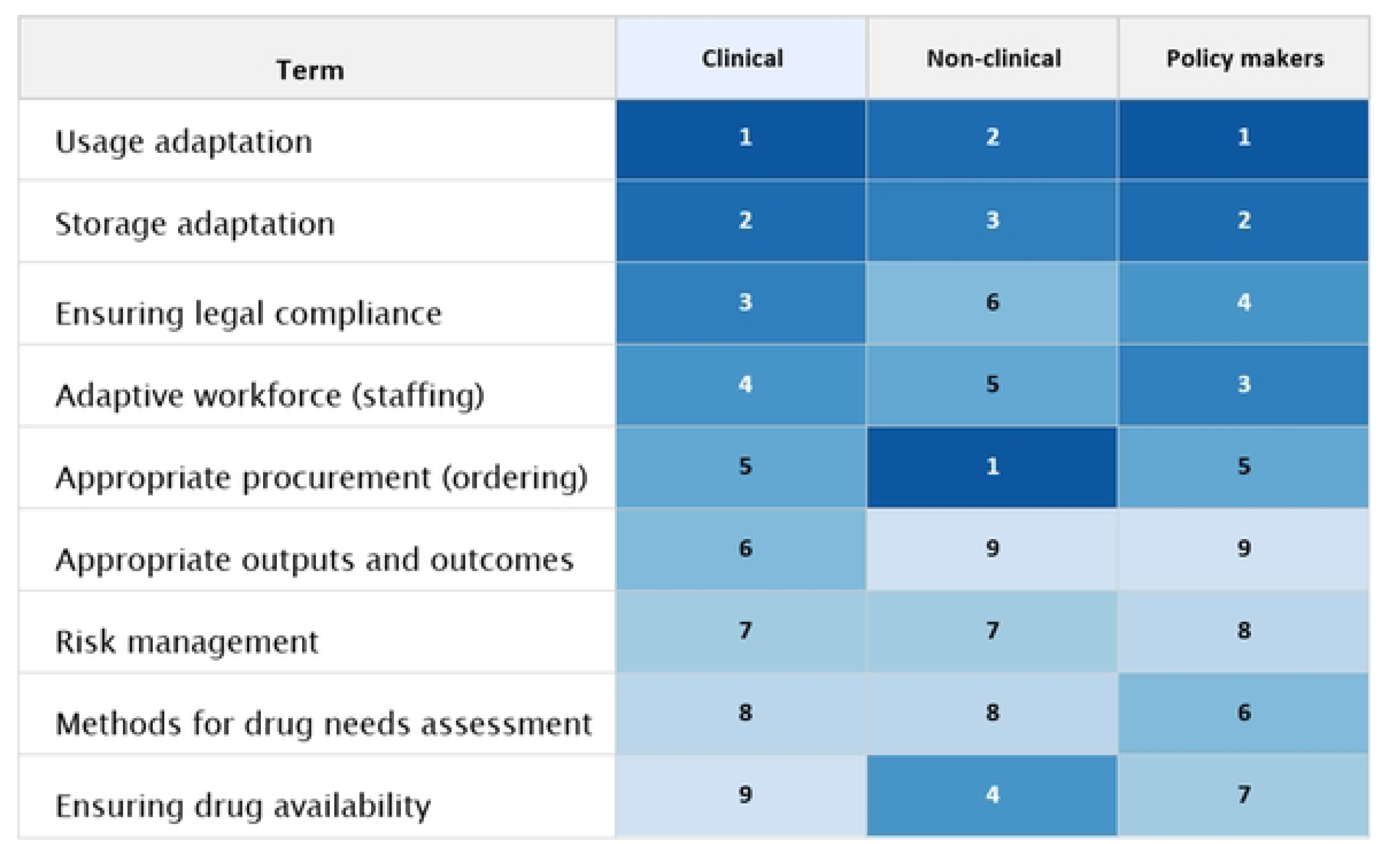
Rank matrix across stakeholder groups. Rows are ordered by the clinical group, progresses from highest priority (rank 1; darkest) to lowest (rank 9; lightest).

In the term of outputs and results: The clinical group placed a higher value on “Appropriate outputs and outcomes” compared to the other two groups. This aligns with the hospital’s direct accountability for treatment outcomes, whereas suppliers and policymakers may focus more on process compliance.

### 3.3. Hierarchy of views by stakeholder group

The ranking of criteria (Q-sort results) varied distinctly across the three groups, reflecting their professional mandates (**Figure 2**). In the views of clinical group, hospital stakeholders focused on operational execution and compliance. Their highest-ranked criteria were Usage (1st), Storage (2nd), and Legal Assurance (3rd). Interestingly, they ranked “Ensuring drug availability” last (9th), possibly indicating that they view availability as a baseline expectation rather than an adaptive variable.

Non-clinical group prioritized logistics and proactive supply management. Their top three criteria were Ordering (1st), Usage (2nd), and Storage (3rd). Conversely, they ranked Risk management and Outputs lowest.

Similar to the clinical group, policymakers prioritized Usage (1st) and Storage (2nd), but placed Human Resources (3rd) higher than other groups. This group ranked Outputs and Results lowest (9th), suggesting a focus on system inputs and processes (HR, Storage) over specific adaptive outcomes in their evaluation model.

### 3.4. Correlation analysis between groups

The Pearson correlation analysis (**Table 2**) quantifies the degree of alignment between the stakeholder groups. There was a strong and statistically significant correlation regarding “Ensuring legal compliance” (NC-CP: r=0.89; NC-PM: r=0.79) and “Storage adaptation” (NC-CP: r=0.73; CP-PM: r=0.88). This confirms that legal compliance and storage standards are universally understood and agreed-upon pillars of the supply chain.

**Table 2.** Correlation values between stakeholder groups Notes: r = Pearson correlation coefficient; p = p-value. NC = non-clinical service providers; CP = clinical service providers; PM = policy makers.

| Criterion | NC-CP<br>r | NC-CP<br>p | NC-PM<br>r | NC-PM<br>p | CP-PM<br>r | CP-PM<br>p |
| --- | --- | --- | --- | --- | --- | --- |
| Adaptive workforce (staffing) | 0.18 | 0.37 | 0.80 | 0.028 | 0.44 | 0.19 |
| Ensuring drug availability | 0.82 | 0.013 | 0.37 | 0.21 | 0.30 | 0.25 |
| Methods for drug needs assessment | 0.14 | 0.32 | 0.38 | 0.103 | 0.81 | 0.000 |
| Ensuring legal compliance | 0.89 | 0.00 | 0.79 | 0.003 | 0.60 | 0.033 |
| Appropriate procurement (ordering) | 0.73 | 0.019 | 0.71 | 0.025 | 0.65 | 0.042 |
| Usage adaptation | 0.68 | 0.004 | 0.58 | 0.015 | 0.71 | 0.002 |
| Storage adaptation | 0.73 | 0.008 | 0.63 | 0.025 | 0.88 | 0.000 |
| Risk management | 0.43 | 0.05 | 0.27 | 0.158 | 0.73 | 0.000 |
| Appropriate outputs and outcomes | 0.46 | 0.019 | 0.09 | 0.356 | 0.41 | 0.034 |

In contrast, the correlation for “Appropriate outputs and outcomes” was notably weak, particularly between the NC and PM (r=0.09, p=0.356). This lack of statistical significance reinforces the finding that different stakeholders have fundamentally different definitions of what constitutes a successful “result” in drug adaptability.

## 4. DISCUSSION

### 4.1. Interpreting consensus

The Role of Inventory and Buffers The high consensus regarding “Usage Adaptation” and “Storage Adaptation” across all stakeholder groups aligns with the fundamental principles of supply chain resilience. Our findings suggest that stakeholders view physical reserves and tangible usage processes as the primary evidence of adaptability. This supports the work of Ding et al., who identified inventory redundancy and facility redundancy as critical components of a resilient hospital supply chain [17]. Similarly, Hart Nibbrig et al. emphasize that redundancy provides necessary “buffers,” such as inventory, while flexibility supports adaptability through planning adjustments [18]. In the Vietnamese context, where supply disruptions can occur, the reliance on storage as a buffer mechanism is a logical, albeit reactive, adaptive strategy.

### 4.2. Analyzing discordance

A significant finding of this study is the discordance regarding “ Appropriate procurement”. Non-clinical providers (suppliers) ranked this as their top priority, whereas clinical providers (hospitals) ranked it significantly lower. This discrepancy likely reflects the suppliers’ shift toward modern supply chain management paradigms, such as Industry 4.0 and 5.0. Alidoost et al. note that Industry 4.0 technologies facilitate accurate forecasting, planning, and traceability [19]. Suppliers, driven by market efficiency, value the proactive nature of “ordering” as a mechanism to manage demand fluctuations. In contrast, hospitals may perceive ordering as an administrative task rather than a strategic adaptive capability. This view is risky; Abdulkadir et al. argue that supply chain performance hinges on understanding customer demand and sharing information on ordering times [20]. The lack of consensus here suggests a need for better integration. As Ghag et al. suggest, moving toward Industry 5.0 requires collaboration between humans and systems to enhance responsiveness [8].

Synthesizing the findings on consensus regarding storage and usage, juxtaposed with the discordance in procurement and outcomes, distinct structural patterns emerge within the supply chain. While operational activities act as a stabilizing core, strategic priorities at the upstream and downstream ends remain fragmented. To visualize these structural dynamics, we propose a multi-level framework illustrating the interplay between stakeholder perspectives (**Figure 3**).

**Figure 3.**
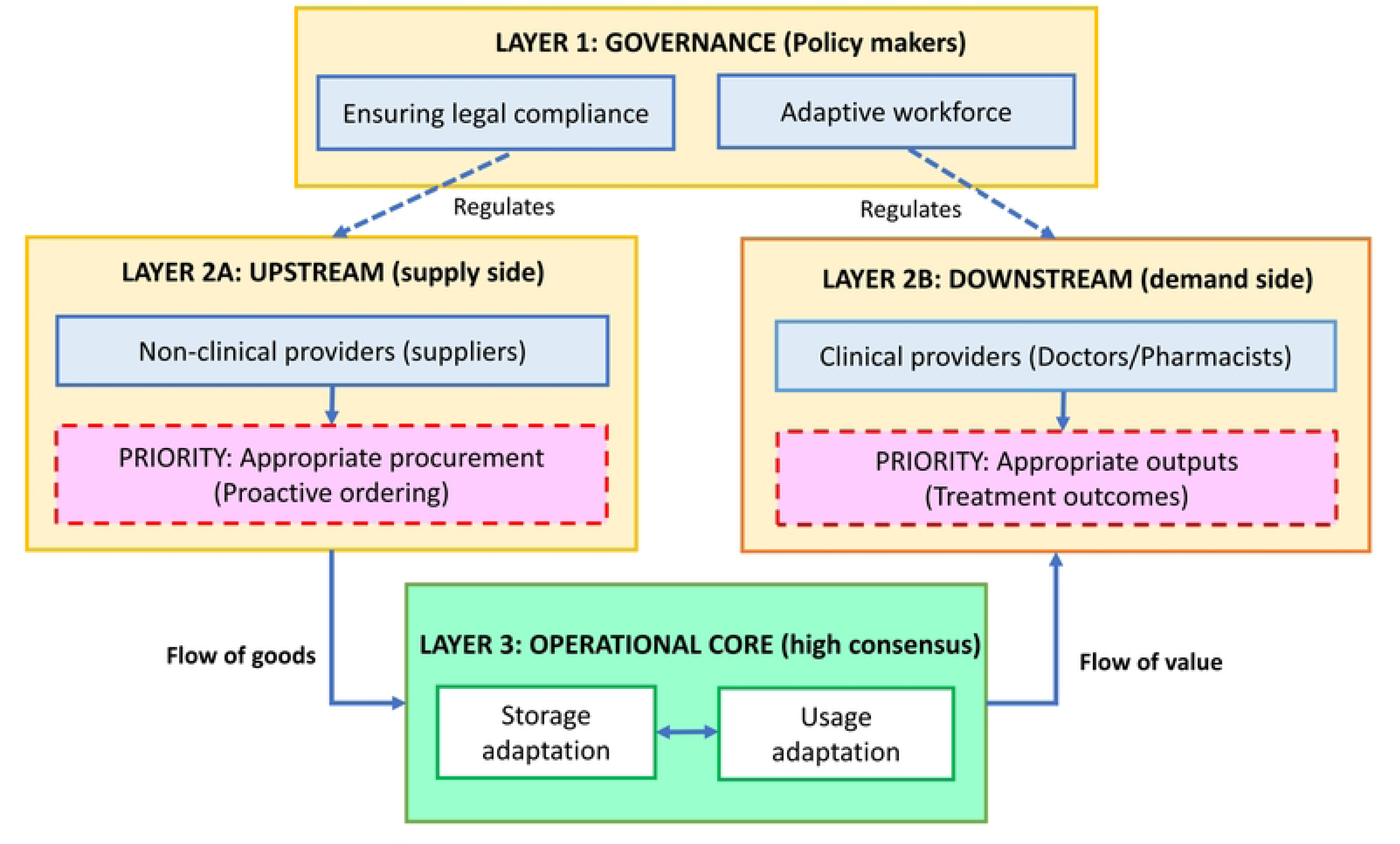
A multi-level framework of stakeholder perspectives on formulary adaptability Note: The framework illustrates the structural interaction of adaptability perspectives. (1) Governance layer: Policymakers establish the regulatory foundation (Legal/HR). (2) Strategic layer (Upstream & Downstream): Shows a discordance where suppliers prioritize proactive *Procurement* (Flow of goods), while clinicians prioritize *Treatment outcomes* (Flow of Value). (3) Operational core: Represents the area of high consensus (*Storage* and *Usage*), acting as the stabilizing anchor for the adaptive system.

### 4.3. The human element in adaptability

While human resources (Adaptive workforce) achieved medium consensus, its importance cannot be overstated. Khan et al. identified “skills and human resource participation” as one of the top three factors impacting pharmaceutical supply chain performance [21]. Our study reinforces this, suggesting that specialized teams are essential for building adaptive capacity. However, the variation in ranking implies that while all agree HR is important, there is no unified standard for how HR should be utilized for adaptability versus routine operations.

### 4.4. Managing complexity and stakeholder perspectives

The healthcare system is a complex adaptive system. The discordance found in our study confirms that different agents (doctors, suppliers, policymakers) operate with different internal rules and goals. Petrie argues that communication between these stakeholders is critical for adaptability [22]. The fact that our study found weak correlation on “Appropriate outputs and outcomes” suggests that stakeholders are currently measuring success differently. Boon et al. recommend evaluation frameworks that are detailed enough for specific cases but general enough to allow for cross-case learning [23]. The indicators developed in this study provide that necessary common language.

### 4.5. Limitations of the study

This study has several limitations. First, the uneven distribution of indicators across the nine criteria may have influenced the ranking results, as criteria with fewer indicators might be skewed by extreme values. Second, the study relies on the perceptions of key opinion leaders rather than direct observation of the supply chain processes. Finally, as the study was conducted exclusively in Vietnam, the findings regarding specific regulatory or operational behaviors may require cautious interpretation when generalized to other healthcare contexts.

## 5. CONCLUSION AND IMPLICATIONS

### 5.1. Conclusion

This study successfully utilized a modified Q-methodology to identify the diverse perspectives of stakeholders regarding drug formulary adaptability. The results indicate a strong consensus on operational adaptability (storage, usage) but significant divergence on strategic adaptability (ordering, results). This confirms that while the physical flow of medicines is well-managed, the strategic alignment between hospital demand and supplier proactivity remains a challenge.

### 5.2. Managerial implications

Based on these findings, we propose three key implications for hospital management and policy:

- Shift to proactive adaptation: Hospitals must elevate “Ordering” from an administrative function to a strategic one, adopting data sharing technologies to align with suppliers’ Industry 4.0 capabilities.
- Standardize adaptive metrics: There is an urgent need to standardize “Appropriate outputs and outcomes” indicators to ensure that clinical, financial, and logistical goals are aligned.
- Human resource development: Investment in specialized supply chain teams is necessary to transition from passive resilience (relying on large inventories) to active adaptability (responsive planning).

By addressing these areas, hospitals can move beyond simple availability assurance to a truly adaptive formulary system that responds effectively to changing disease patterns.

## Data Availability

All relevant data are within the manuscript and its Supporting Information files.

## Supporting information

S1 Appendix.

Initial indicator set and expert consultation form (Round 1). (S1_Appendix.PDF)

S2 Appendix.

Refined indicator set and re-evaluation form (Round 2). (S2_Appendix.PDF)

## REFERENCES

1. Yaroson EV, Breen L, Hou J, Sowter J. Advancing the understanding of pharmaceutical supply chain resilience using complex adaptive system (CAS) theory. Supply Chain Management: An International Journal. 2021;26(3):323–340. doi:10.1108/SCM-05-2019-0184

2. Goldberg RB. Managing the Pharmacy Benefit: The Formulary System. JMCP. 2020;26(4):341–349. doi:10.18553/jmcp.2020.26.4.341a

3. Gurupur VP. A review on advances in design and development of complex adaptive systems for healthcare using concept maps. Health Technology. 2021;5(0). doi:10.21037/ht-21-12

4. Franco-Trigo L, Fernandez-Llimos F, Martínez-Martínez F, Benrimoj SI, Sabater-Hernández D. Stakeholder analysis in health innovation planning processes: A systematic scoping review. Health Policy. 2020;124(10):1083–1099. doi:10.1016/j.healthpol.2020.06.012

5. Olazabal M, Galarraga I, Ford J, Sainz De Murieta E, Lesnikowski A. Are local climate adaptation policies credible? A conceptual and operational assessment framework. International Journal of Urban Sustainable Development. 2019;11(3):277–296. doi:10.1080/19463138.2019.1583234

6. Marrone PV, Mathias FR, Bernardo WM, et al. Decision Criteria for Partial Nationalization of Pharmaceutical Supply Chain: A Scoping Review. Economies. 2023;11(1):25. doi:10.3390/economies11010025

7. Marube N, Longaray AA, Ensslin L, Ensslin SR, Dutra A. EVALUATING SUPPLY CHAIN MANAGEMENT PERFORMANCE IN PUBLIC HEALTH CARE: AN MCDA APPROACH. Pesqui Oper. 2024;44:e275847. 10.1590/0101-7438.2023.043.00275847

8. Ghag N, Sonar H, Sawant R, Bankapalli K. Examining industry 5.0 and supply chain performance: an empirical study for managing supply chain complexity. Production & Manufacturing Research. 2025;13(1):2501181. doi:10.1080/21693277.2025.2501181

9. Kazakov R, Howick S, Morton A. Managing complex adaptive systems: A resource/agent qualitative modelling perspective. European Journal of Operational Research. 2021;290(1):386–400. doi:10.1016/j.ejor.2020.08.007

10. Bastani P, Dehghan Z, Kashfi SM, et al. Strategies to improve pharmaceutical supply chain resilience under politico-economic sanctions: the case of Iran. Journal of Pharmaceutical Policy and Practice. 2021;14(1):56. doi:10.1186/s40545-021-00341-8

11. Apeh CE, Odionu CS, Bristol-Alagbariya B, Okon R, Austin-Gabriel B. Reviewing healthcare supply chain management: Strategies for enhancing efficiency and resilience. IJMRGE. 2024;5(1):1209–1216. doi:10.54660/.IJMRGE.2024.5.1.1209-1216

12. Yule EL, Donovan K, Graham J. The challenges of implementing adaptation actions in Scotland’s public sector. Climate Services. 2023;32:100412. doi:10.1016/j.cliser.2023.100412

13. Ansah EW, Amoadu M, Obeng P, Sarfo JO. Health systems response to climate change adaptation: a scoping review of global evidence. BMC Public Health. 2024;24(1):2015. doi:10.1186/s12889-024-19459-w

14. Dinh TS, Brueckle MS, González-González AI, et al. Stakeholder Perspectives on the Development and Implementation of a Polypharmacy Management Program in Germany: Results of a Qualitative Study. J Pers Med. 2022;12(7):1115. doi:10.3390/jpm12071115

15. Fritz MMC, Rauter R, Baumgartner RJ, Dentchev N. A supply chain perspective of stakeholder identification as a tool for responsible policy and decision-making. Environmental Science & Policy. 2018;81:63–76. doi:10.1016/j.envsci.2017.12.011

16. Saesen R, Lejeune S, Quaglio G, Lacombe D, Huys I. Views of European Drug Development Stakeholders on Treatment Optimization and Its Potential for Use in Decision-Making. Front Pharmacol. 2020;11. doi:10.3389/fphar.2020.00043

17. Ding B, Yang X, Gao T, Liu Z, Sun Q. Confirmation of a measurement model for hospital supply chain resilience. Front Public Health. 2024;12:1369391. doi:10.3389/fpubh.2024.1369391

18. Hart Nibbrig M, Sharif Azadeh S, Maknoon MY. Adaptive resilience strategies for supply chain networks against disruptions. Transportation Research Part E: Logistics and Transportation Review. 2025;200:104172. doi:10.1016/j.tre.2025.104172

19. Alidoost F, Mustafee N, Monks T, Harper A. Simulation in healthcare supply chains with perishable products: a scoping review. Journal of the Operational Research Society. Published online June 2, 2025:1–36. doi:10.1080/01605682.2025.2509698

20. Abdulkadir R, Matellini DB, Jenkinson ID, Pyne R, Nguyen TT. Assessing performance using maturity model: a multiple case study of public health supply chains in Nigeria. Journal of Humanitarian Logistics and Supply Chain Management. 2023;14(1):17–70. doi:10.1108/JHLSCM-05-2022-0053

21. Khan SA, Gupta H, Gunasekaran A, Mubarik MS, Lawal J. A hybrid multi-criteria decision-making approach to evaluate interrelationships and impacts of supply chain performance factors on pharmaceutical industry. Journal of Multi-Criteria Decision Analysis. 2023;30(1-2):62–90. doi:10.1002/mcda.1800

22. Petrie DA. Integration as innovation in healthcare systems. Healthc Manage Forum. 2025;38(2):76–83. doi:10.1177/08404704241292629

23. Boon E, Body NS, Biesbroek R. Developing and testing an evaluation framework for climate services for adaptation. Climate Services. 2025;38:100549. doi:10.1016/j.cliser.2025.100549

